# Accelerating precision medicine: a proposed framework for large-scale multiomics data integrity, interoperability, analysis, and collaboration in biomedical discovery

**DOI:** 10.1101/2024.03.15.24304358

**Authors:** Vivek Sriram, Ashley Mae Conard, Ilyana Rosenberg, Dokyoon Kim, Amanda K Hall

**Author notes:** **Correspondence:** Amanda K. Hall, PhD **Postal Address:** Microsoft Building 99, 14820 NE 36th Street, Redmond, Washington, 98052, USA **Email:**.

## Abstract

**Objective:** To identify and define a process and framework for biomedical discovery research. Our study aim was to characterize the biomedical discovery lifecycle across data modalities and professional stakeholders involved in biomedical research to address the multiomics data challenges of precision medicine.

**Materials and Methods:** We recruited fifteen professionals from various biomedical roles and industries to participate in 60-minute semi-structured interviews, which involved an assessment of common challenges, needs, tasks, and data management methods and a brainstorm exercise to validate each professional’s biomedical research process. We applied a qualitative analysis of individual interviews using a constant comparative approach for emerging themes.

**Results:** We found a general process of biomedical discovery across all participants that consisted of four key stages: data plan, data integrity, data analysis, and data-driven discovery. Within each stage of the process, participants highlighted their challenges and needs and emphasized the importance of data integrity and interoperability, particularly during data hand-offs. The process extends across three general levels of data, including non-human, non-clinical human, and clinical human data, to define an overarching framework for biomedical discovery research.

**Conclusions:** The proposed framework provides researchers with an opportunity to align workstreams and converge on a single process to conduct biomedical research across stakeholders and data types. Key opportunities were found that can be explored in the health technology space, including generative artificial intelligence, to help tackle multiomics data challenges to advance precision medicine.

## INTRODUCTION

Fulfilling the promise of precision medicine necessitates a standard process for biomedical discovery given the diversity of data and stakeholders involved. *Biomedical discovery* involves the investigation of disease etiology and the elucidation of underlying mechanisms of biological processes. *Precision medicine* aims to achieve a more accurate and precise version of medicine that uses large-scale, multi-modal data to characterize the underlying mechanisms of disease onset across cohorts of patients and improve outcomes in clinical settings. The ultimate goal of precision medicine is to transform patient care through individualized disease prediction, prevention, treatment, and therapeutics.[1, 2]

The currency of both precision medicine and biomedical discovery has always been data. Precision medicine begins with the integration of *multiomics datasets*, data that correspond to different levels of biological structure[3], for the purpose of gaining a comprehensive understanding of human health. These insights can assist healthcare professionals in personalizing patients’ diagnoses and treatment.[4]

The advent of *big data*, the exponential increase in variety and quantity of data that are collected, has significantly disrupted the field of biomedical discovery, leading to a rapid increase in the pace of innovation.[5] In only the past few years, big data have facilitated the complete sequencing of the human genome[6], pioneered chimeric antigen receptor (CAR) T-cell therapy for cancer[7], and contributed to the development of novel vaccines during the COVID-19 pandemic[8].

Despite the many advances that were made across the field of biomedical discovery, a lack of data quality and interoperability has left the ultimate promise of precision medicine unfulfilled. Therapeutic medicine has remained largely unchanged over the past twenty years, with minimal benefits to public health and ever-expanding research and development costs.[2] This gap between biomedical expectations and reality ultimately arises from inaccurate assumptions regarding the integrity and utility of the data. Indeed, most efforts to improve the pace of biomedical research focus on advanced tooling while incorrectly assuming the quality and integrity in the data.[9] However, no amount of technical sophistication can generate meaningful insights from substandard data.[10]

The failure to achieve an improved version of medicine is not just a disappointment – it is unequivocally harmful. If we do not fix the current trajectory of precision medicine research, then the many existing health disparities within our medical systems will persist and worsen as more biased, substandard biomedical data continue to accumulate. Accelerating the pace of biomedical discovery through a renewed focus on data quality and integrity is essential to ensure the populations in our health systems that are already underserved are not left behind.

There is an opportunity to establish a unified approach to research across the landscape of biomedical discovery to achieve our target of enhanced data integrity. Although contextual nuances vary extensively, we can identify a consistent set of problems related to data integrity across subdisciplines of biomedical discovery research. In the following sections, we highlight individual issues faced by research involving the following biomedical fields: life science, genomics, epigenomics, metabolomics, immunology, clinical research, and multiomics. For instance, life sciences research experiences challenges with data scale and heterogeneity, isolated resources, and insufficient compute. Genomics research faces problems with dataset isolation, as well as a lack of scalability and a lack of trust in data. Epigenomics research must account for experimental complexity and analysis standardization.

Metabolomics research encounters significant problems regarding data reproducibility, while immunology research confronts a lack of metadata standardization. Clinical research must maneuver a muddled regulatory environment, and finally, multiomics research must handle multimodal data integration. Other examples of relevant biomedical data modalities for which we do not provide further detail include transcriptomics, proteomics, single cell ‘omics, and image-based data.[11] Within each biomedical subdiscipline we cover, we identify known challenges and discuss potential options to address these issues and improve the efficiency of research. We then take the challenges and needs identified in each subdiscipline to converge upon a common set of data integrity issues that must be addressed to accelerate biomedical discovery and advance precision medicine.

### Life Science

A variety of complex computational tasks impact life science research. For instance, large-scale datasets and heterogeneous data formats are common, and there is a lack of tooling that can simplify their exploration and integration.[12] Furthermore, local data are often siloed within disparate institutions due to differences in collection protocols and inadequate computational infrastructure.[12] These challenges, coupled with a lack of sharing incentives in the field, result in redundant efforts and wasted resources. Data analysis has also become more laborious, requiring sufficient compute to make data mining and the development of complex biological models and digital twins possible.[12]

No longer is it feasible or practical to continue advancing progress in core biological research without an adequate informatics infrastructure.[12] Cloud-based platforms play a vital role in translating basic biological research into knowledge-based innovations. Cloud compute is essential to enable convenient access to shared data, resources, and scalable informatics tooling.[12] Along with these technology-based requirements, a need for improved data accessibility and collaboration is likewise apparent.

Collaborative community partnerships between federal organizations, research institutions, and industrial partners will be essential to decide on data access protocols and sharing standards, as well as tool evaluation and distribution within these cloud-based platforms.[12] Finally, open-source cooperation and innovation must become indispensable if the field of life sciences is to be democratized.

### Genomics

Numerous technical considerations have diminished the impact of genomics-based research, particularly with respect to clinical implementation. Similar to other biomedical data modalities, genomic data are complex, high-dimensional, and often separated across different institutions.[13] Depending on the type of data being analyzed, experimental complexity, scalability, and restrictive costs can also hinder progress. In the case of rare genetic disorders, the segregation of genetic datasets can pose challenges in the identification of patient cohorts for research, particularly for phenotypes with low penetrance.

Furthermore, genomic data that are not properly de-identified can implicate not only individuals but also their relatives, ancestors, and descendants.[14] Although many associations between genetic variants and disease phenotypes have been identified, pinpointing the genetic components responsible for diseases remains a significant task. Research has demonstrated that most genetic associations for traits involve the combined effect of multiple small-effect variants working together rather than a single genetic target which could implicate the disease.[15] Identifying other contributors to missing heritability in these scenarios, as well as the extra steps that must be taken through experiments such as mendelian randomization to ascertain genetic causality, can further complicate researchers’ abilities to identify proper genetic targets for novel therapeutics. Clinicians often lack the necessary expertise to interpret the nuances of genetic variants associated with a disease, requiring collaboration with clinical experts and introducing latency to discovery efforts. Even with the participation of specialists, it remains challenging to determine the usefulness of in-depth, comprehensive genomic testing for patient treatment, as there is no clear standard for evaluating its benefit.[16] Overall, a lack of trust in genomic data is the largest obstacle to the implementation of genomic medicine.[14] Currently, there is limited visibility regarding data storage and access management, the purpose of different datasets, and the validity of discoveries made from these data.

### Epigenomics

Epigenomics research shares many of the same challenges as other ‘omics fields, as well as its own set of technical challenges unique to the data modality. *Epigenetics* and *epigenomics* research refer to the study of phenotypic changes based upon alternations to genetic organization.[20] Epigenomics offers a promising approach to uncover much of the heritability that has been missed through standard genomics perspectives, suggesting new therapies for the treatment of various diseases. Challenges common to other data modalities include the creation of diverse sample cohorts for research, the assurance of patient privacy, and the prevention of patient de-identification.[17] Furthermore, epigenomics researchers often have to collect both longitudinal and tissue-specific data to be able to study the dynamism of genetic organization.[17] These two facets add complexity to both experimental design and computational analysis. The specific genetic and metabolic features that contribute to epigenetic regulation also vary drastically from one another. As a result, there is extensive inconsistency and confusion regarding the ground truth thresholds that are required to mark the presence of a given feature in the data.[17]

### Metabolomics

Research in *metabolomics*, involving the comprehensive analysis of biomolecules and metabolites to provide profiles into the state of patients’ biological processes and metabolic pathways, faces many of the same issues as experimentation in other data modalities.[18] Similar to other sequencing-based subdisciplines, metabolomics research often includes challenges such as read quality, depth, and partial coverage.[3] Another significant challenge in the field is that different biomolecules can have completely different characteristics from one another, making it difficult to quantify the relative significance of each metabolite.[3] Furthermore, a substantial amount of technological variation exists across institutions.

The quality of reagents is known to vary widely, with false signals often arising from experimentation.[19] The general distrust of data accuracy and the inability to present gold standards for metabolomic research has hindered the translation of preclinical models into the clinic. Indeed, small errors in the data or in upstream analysis can lead to significant failures in downstream clinical trials.[19] To enhance reproducibility and trust in biomedical discoveries taken from metabolomic data, common quality control and analysis pipelines must be agreed upon across the field.

### Immunology

*Immunology* involves the study of immunological components, including genes and molecules, to gain a deeper understanding of the overall workings of the immune system. However, while data in this field continue to grow, analysis has fallen behind due to challenges including scale, high dimensionality, and heterogeneity across institutions.[20, 21] Approaches such as manual gating for data visualization and clustering require additional time and effort from experts. Finally, there is an apathy in the discipline with respect to data sharing. Both the research and clinical communities tend to generate novel data instead of repurposing shared data, resulting in duplicated research and wasted time and effort.[21]

### Clinical Data

Among the various biomedical data modalities, clinical data and the drug discovery process are arguably the most well-documented and analyzed. A variety of regulatory and statistical models exist that facilitate clinical research, and the pharmaceutical industry at large has spent billions of dollars to define and accelerate the drug development pipeline. Despite these investments, however, several significant challenges still exist in the use of clinical data and clinical trials for drug discovery, development, and validation. Given the large-scale, longitudinal nature of clinical discovery, healthcare and research institutions often have difficulty recruiting appropriate, representative cohorts for their studies[11], as well as ensuring retention throughout the course of the trial[22]. Furthermore, new technologies and therapies have led to additional complications of trial design, study complexity, and monetary expenses.[23] Clinical research efforts often involve collaboration among multiple institutions, necessitating effective communication among the stakeholders planning the trials, the clinicians generating the data, and the researchers analyzing the data.[24] However, there is often insufficient coordination among these parties, and clinical research informatics platforms struggle to integrate with existing workflows. Unstructured clinical data can also be challenging to process[11], and a variety of additional factors, such as the type of research, the clinical setting, desired endpoints, the development phase of the drug, and the efficacy of data capture, can influence the ultimate utility of data that are collected.[25] Researchers need to appropriately handle confounders not represented in the data to gain an accurate representation of association and causality[26]. With much of clinical research focused on data derived from electronic health records, biases that arise due to prior clinical practices as well as the focus on billing over research must also be considered.[5, 27] Finally, the regulatory environment for clinical research and drug development remains muddled. In consideration of the development of new machine learning technologies, therapeutics, and larger patient datasets, the clinical research community would benefit from established well-designed standards to prioritize equitable patient safety and privacy.[24]

### Multiomics

As research and the clinic move toward more comprehensive views of biomedical systems, multiomics approaches that can consider more than one data modality at a time have become increasingly crucial. Nevertheless, despite a plethora of technical advancements, progress in multiomics research continues to fall behind other fields. Indeed, multiomics research amplifies the issues that are seen with individual data modalities – regardless of analytical ability, problems with data cause problems in the results.[10] Bias and noise are particularly prominent in clinical settings, and data are often not representative of the populations they are intended to represent.[5, 27] Due to concerns regarding patient privacy as well as a culture of competition across research groups, segments of data are typically restricted into isolated systems rather than in a common data storage. This separation of data reduces their impact, limiting researchers’ ability to characterize the full state of a patient’s health.[28] Data collection can have significant influences downstream, as an adequate sample size is needed to ensure that knowledge-based discoveries have sufficient statistical power.[29] Even after collecting reasonable data, researchers still must apply quality control (QC) processes to restrict variability in analysis. However, there is currently limited availability of QC schemes for many biomedical research groups working with multiomics data.[30] Apart from genomic data, which have the standardized GATK toolkit for primary and secondary analysis, discovery involving other data modalities has no clear reference procedures that can determine the clinical utility of biomarkers and other molecular phenotypes.[30] Issues such as budgetary constraints and privacy concerns lead to decreased data harmonization across institutions, resulting in lower reliability of results.[30] Data integration is another significant challenge in multiomics analysis. With the diverse signal and noise of different data modalities, increased compute and expert biological interpretation are often needed but often lacking. ML-based data standardization for multimodal integration is still highly context-dependent – despite their extensive use in research environments, ML models are currently unable to influence translational applications of multiomics research. This discrepancy compared to ML’s success in other fields stems from the fact that such methods were not intended for medicine in the first place.[31] ML learns from prior data, meaning that it can reflect all biases represented in a system, however there is an inherent tradeoff between a ML model’s explainability and its accuracy.[31] As a result, the development of multiomics-focused ML models requires insight from clinical experts throughout the research process.[11] This requirement is a significant bottleneck in the broad applicability of current learning health systems, Finally, many research centers possess insufficient compute capabilities to successfully develop their own computational models for multiomics research.

Given the challenges identified for individual ‘omic subtypes as well as multiomics modeling, multiple common issues related to precision medicine research emerge. Across all subdisciplines, researchers are handling large-scale, complex, high-dimensional data that include a variety of heterogeneous formats.[32] These data are typically isolated within their respective institutions, hindering efforts toward reproducibility, and prevent efforts to generate diverse, longitudinal, comprehensive patient cohorts.

A variety of stakeholders are involved in biomedical discovery and precision medicine research, including healthcare systems, clinical laboratories, technology companies, academia, and the government.[33] The promise of precision medicine and the development of accurate biomedical digital twins rely on the ability of these stakeholders to collaborate with one another and link diverse, high-quality data across ‘omic subtypes. Without a standard agreed workstream to process and validate data collected across a multitude of studies, the output of biomedical data will not be as usable to new knowledge discovery. Efforts to improve tooling and modeling all assume data quality.[9] A lack of interoperability standards is particularly problematic for production-level models in healthcare and life science applications, where poor data quality could cause erroneous results and ultimately harm to patients.

Each biomedical subdiscipline assumes that its work differs from the rest, however if we could identify similarities across data modalities and converge on a single process for biomedical discovery research could we drastically reduce the time required to develop an individualized understanding of disease. Only through participation from stakeholders across basic sciences, translational research, clinical, and public health can we hope to reach a unified process to deliver population-level health benefits.

### Study Objectives

Given the importance of data quality and interoperability in biomedical research and the challenges that continue to hinder personalized medicine, the aim of our study was to better understand the overarching process of biomedical discovery research across stakeholders and biomedical data types. Our objectives are to (1) identify and define the processes and tasks performed by biomedical researchers, (2) evaluate researchers’ needs and challenges related to data, data management, and collaboration, and (3) assess the analytical tools and workflows that researchers leverage to conduct their work. We aim to explore a framework for the process of biomedical discovery research.

## MATERIALS AND METHODS

We conducted fifteen sixty-minute semi-structured interviews with individuals placed throughout the field of biomedical discovery, including computational biologists, research scientists, data curators, data stewards, and data generators. The first part of each interview focused on the participant’s background, research objective, general tasks and jobs-to-be-done, data and tooling needs, and current challenges. The second part of each interview focused on a brainstorming exercise. We present details on participant recruitment, informed consent process, data collection and analysis methods below.

### Ethics Statement and Participant Recruitment

We conducted our study with fifteen professionals who work in biomedical discovery research in the United States (US). Our study criteria consisted of participants of age range 18 to 100, who work in biomedical discovery in the US, and speak English. Our study (protocol ID 10415) was reviewed and approved by Microsoft Research Institutional Review Board (IRB). Written informed consent was obtained from each participant prior to the start of the interviews.

Participants were recruited through a research recruitment company that recruits for research studies across the US. Participants were recruited through a combination of methods such as active outreach and internal study panel contact databases. A detailed participant screener was applied, and pre-approval was performed by the research team. Pre-approved and interested participants who met study eligibility criteria were informed about the purpose of the study and provided a copy of the informed consent. Interested participants who provided written informed consent to the research recruitment company were then scheduled for an interview. Prior to the start of each interview session, participants were asked if they had any questions related to the study and confirmed they had read and signed the informed consent. Participants were compensated $175 USD for their time via a gift card distributed through the research recruitment company.

### Data Collection

Participants were asked questions related to their professional roles, the type of work they conduct, the research problems they are trying to solve, the data and tools they use, their challenges and needs, and to relate their day-to-day research tasks in the first half of the interview. In the second half of the interview, the research team displayed the research diagram (Figure 1) on their screen and asked questions related to how similar or different the diagram flow was to the participants’ research processes, where in the diagram flow they position their day-to-day roles, and what information was amiss as well as suggestions for how to accurately represent each stage of their research process. Figma (https://www.figma.com) was used for the virtual whiteboard brainstorm portion of the interview and sticky notes were used to capture participants feedback in real-time to allow them to clarify and validate their research process. Figure 2 depicts an example of the notetaking process. At the end of each interview, participants were asked general quantitative demographic questions.

**Figure 1.**
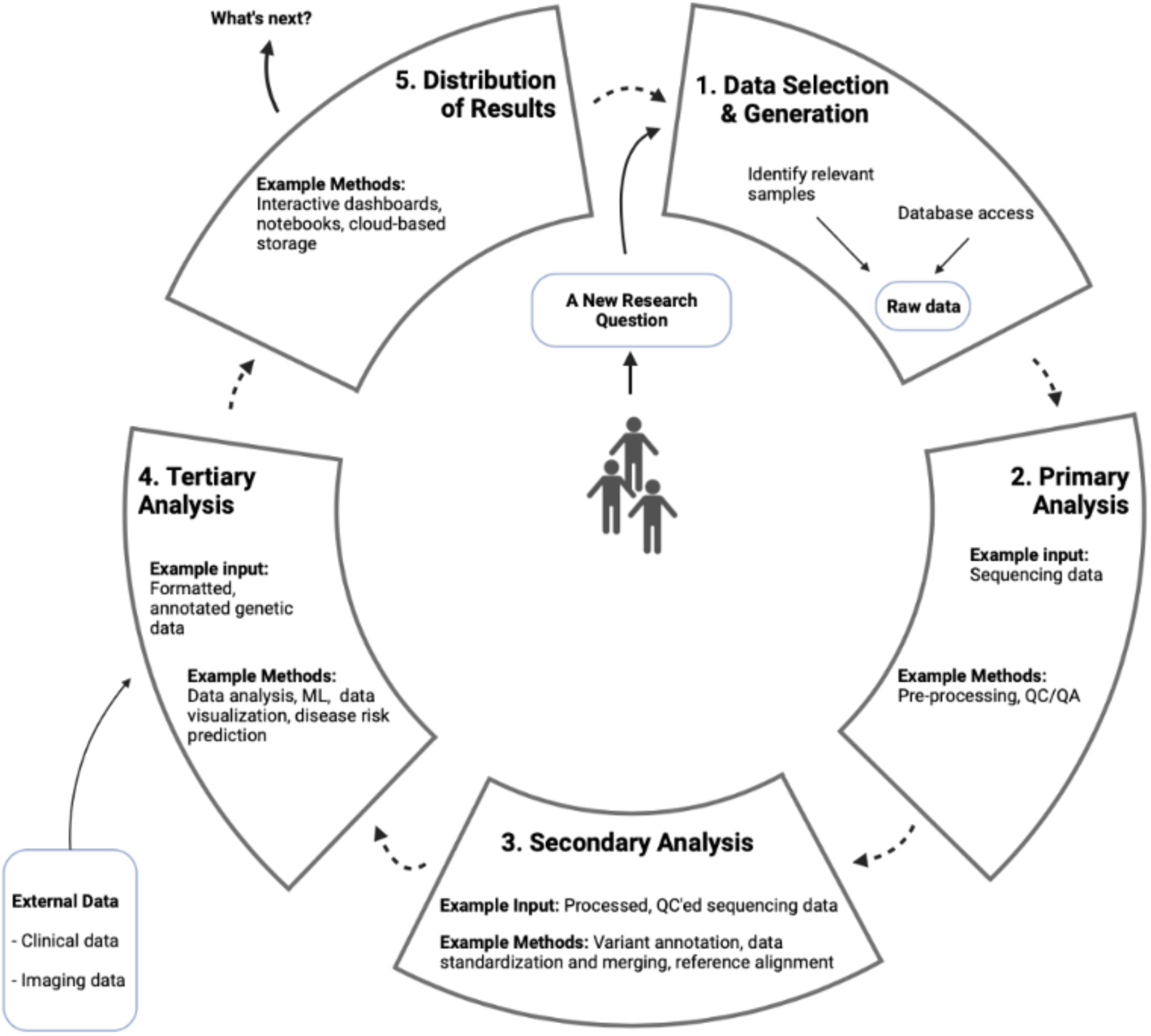
Baseline visualization for brainstorming exercise. Created with Biorender.com

**Figure 2.**
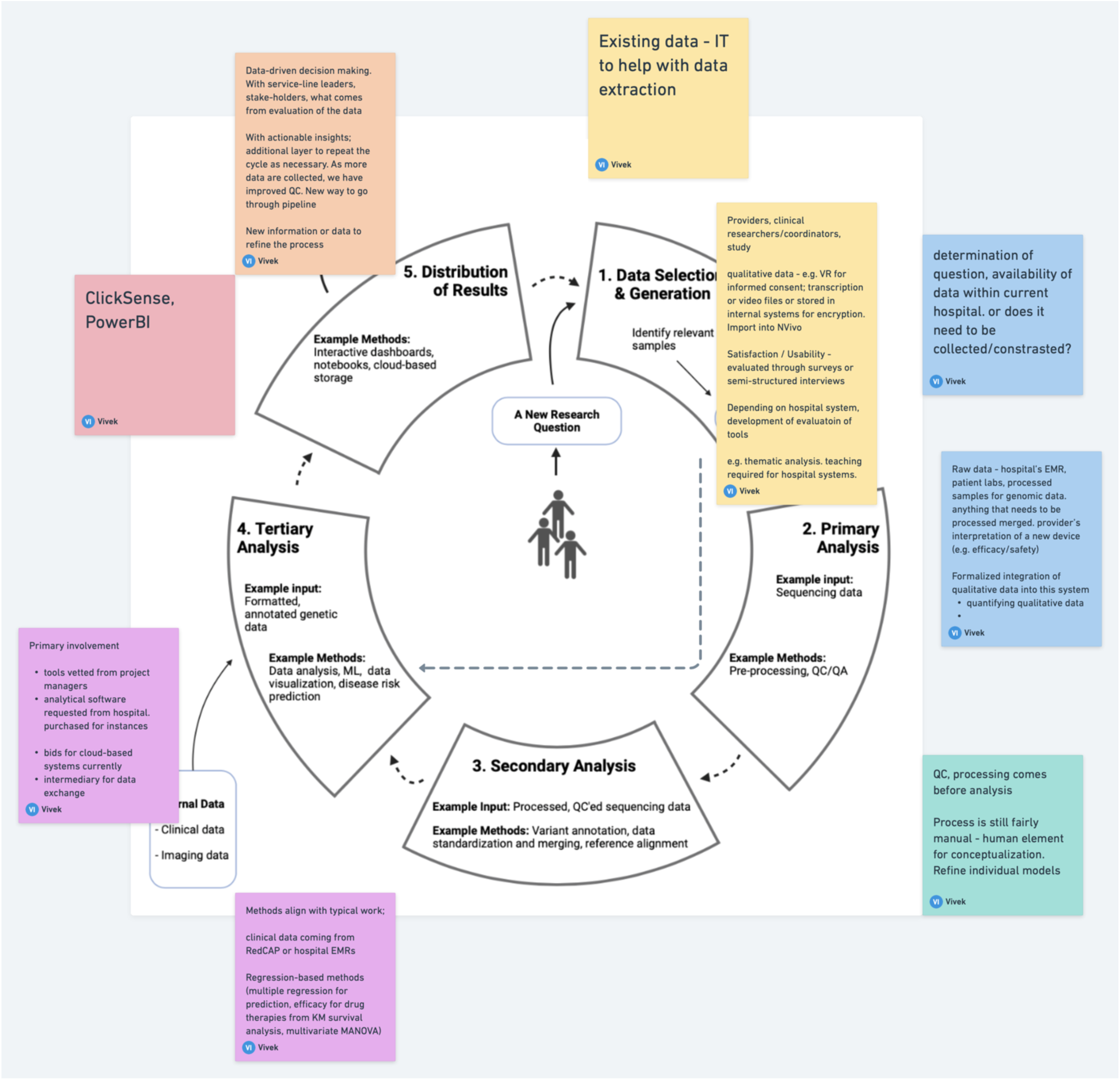
Example of the note-taking process during the brainstorm exercise.

We created the diagram (Figure 1) as a brainstorm tool to elicit feedback from participants during the interviews to validate their research process [34, 35]. We searched the literature to identify a pre-existing example for a research process visual to leverage for the brainstorm exercise but could not find anything sufficient based on our study objectives in the published literature. Therefore, we drafted a diagram based on standard genomics data processing and analysis [36].

### Data Analysis

All interviews were conducted via Microsoft Teams video conference platform from July to August of 2022 by the first author (VS) and audio-recorded with participants’ informed consent. Microsoft Teams auto transcription was used and then each interview transcript was verified and corrected for accuracy later via the recordings by authors VS and AKH. The interviews lasted 0:47:33 to 1:13:19 minutes. The first and last authors met periodically to discuss interviews and identify emerging themes. We used a combination of inductive and deductive thematic coding approaches to the qualitative data.[37] Initial themes consisted of ‘data collaboration’ and ‘data quality’ and phases of analysis.’ As the interviews progressed, we iterated over the data to produce higher-level themes, including ‘data extraction’ and ‘access’, ‘clinical trial data platforms’, ‘analysis processes’, and ‘data hand-offs’. Finally, we abstracted out the main themes that became the main stages of the biomedical discovery process, ‘data plan’, ‘data integrity’, ‘data analysis’, and ‘data-driven discovery’, and ultimately the final framework with the additions of ‘level A’, ‘level B’, and ‘level C’ to represent the various biomedical data modalities and applications used across the stages.

## RESULTS

### Quantitative Findings

All participants lived in the US, worked in biomedical discovery research, and worked with a range of nonclinical, clinical, imaging, and genomics data. The age range of participants were 18-24 (1), 25-34 (8), 35-44 (4), and 45-54 (2). Their work experience ranged from 1-5 years (5), 5-10 years (4), and more than 10 years (6). Our study included 5 females and 10 males. Most participants identified as Caucasian/European descent (9), followed by South Asian (3), East Asian (1), African Descent (1), and other/Mixed Ancestry (1). Participants worked in a variety of industry and academic settings ranging in size from self-employed freelance positions to companies with over 20,000 employees, with about half from pharmaceutical or biotechnology operations and the other half from academic medical centers, healthcare organizations, or hospitals.

### Qualitative Findings

Analysis tools were highly context-dependent, participants used IBM SPSS, REDCap, and Microsoft Excel for intuitive computation, ImageJ and Prism for image analysis, GATK for primary and secondary genomic data, Python (including pandas, NumPy, SciPy packages), R (including Bioconductor, ggplot, tidyverse libraries), SQL, and SAS for general data needs, Nextflow and Cromwell for pipelining and workflow development, and Anaconda and Docker for versioning of software environments. The most common research motivations that participants discussed were the development of new domain-specific insights, to identify cohorts for clinical trials, accelerate drug development, bring therapeutics to patients, facilitate FDA regulatory approval, simplify patient diagnosis, and discover positive changes that could be implemented in clinical settings for improved patient health outcomes.

Four main themes emerged from the data analysis of participant interviews based on the data journey stages of biomedical discovery research: Data Plan, Data Integrity, Data Analysis, and Data-Driven Discovery. Below are the aggregated findings along with the main challenges and needs of participants by each stage of participants’ biomedical research process.

## 1. Data Plan

A primary focus across participant interviews is the balance between identifying and extracting the appropriate data for a given research question. Participants described a variety of data types with which they worked (Supplemental Table 2), including protein abundances from model organisms, structured and free-text clinical data, genomic single-cell and whole genome sequencing data, and post-clinical data, such as drug performance and marketing metrics. This process to identify the ideal data for a research question constitutes the first main theme: “Data Plan.”

Regardless of the data modalities under consideration, participants expressed many of the same complaints with respect to their data. A key challenge regarding work in the Data Plan was the tradeoff between data generation and data extraction. Both sufficient financial resources and an adequate amount of time were found as needs to create the data or to procure it from an external source. Paper-based data collection was described as a tedious manual process for much of wet lab data generation, leading to increased risk of downstream quality issues when transferring data into computational environments. Complications regarding coordination and collaboration among stakeholders and research planners to identify suitable data were also cited as a common issue at the start of biomedical research.

Specific needs described by participants for this phase of research included more trustworthy and simplified automated processes for electronic wet lab data collection, secure systems for the storage of big data, the ability to store multiple file types along with options for custom organization and identification, more computationally efficient methods for data transfer, and tools to facilitate quicker, intuitive data warehousing and withdrawal. P05 explained the significance of efficient data access as follows:

> *“ In drug discovery, … being able to pull up data from past experiments is really, really critical when you are working on documents … [because] at any point someone from the FDA could say, “… [I want to] see the data from this experiment that you’re referencing in this, you know, new drug application.” And you need to be able to put your hands on it pretty quickly, and it needs to be in a nice tight package. It needs to be searchable.”* – (P05, biotechnology consultant)

The raw, non-curated data that leave the Data Plan experience a handoff among personas including data collectors and custodians, lab technicians, and research scientists as they continue through the biomedical data lifecycle. During these data transfers, common challenges for participants included a lack of unity among data management and sharing systems, prohibitive data storage costs, and complicated coordination among multiple stakeholders.

## 2. Data Integrity

The second theme identified from our interviews was the stage of “Data Integrity.” During this period, data are cleaned and reformatted for use in analysis. This work is often performed by a data scientist or a data curator, with oversight from a data management entity. In the case of ‘omics data, best practice workflows for primary and secondary analysis such as the GATK are applied. The numerous methods described by participants for storing (Supplemental Table 3), sharing (Supplemental Table 4), and managing access (Supplemental Table 5) highlight the significance of data movement during the Data Integrity step.

Common challenges described by participants for this period included the need for expert involvement to evaluate data accuracy and security, a lack of consistency in requirements for quality control procedures, compliance with regulatory expectations, extensive interaction across stakeholders to ensure data are handled properly, and lag time during data curation, particularly when processing unstructured data. P13 described the issues that arose when he was recruited to organize electronic health record data for a medical center:

> *“Originally when I was hired on, this clinic was using their EMR just as like “Hey, just get the data in there. Who cares how it’s organized … Leave it to … the people who we’re sending it to … make sense of it all,” which isn’t a very nice thing to do! So … one of the first things that I was tasked with … was organizing their workflow and how they’re entering data and get[ting] the correct tools in place and utiliz[ing] their EMR properly so that they’re putting in actual discrete data points… [T]he EMR had that capability, they just weren’t using it that way… [T]hey were entering in medications, allergies, CPT codes…, original ordering provider diagnosis, all of this stuff in … just basically a single block of text.”* – (P13, clinical data curator)

Tedious data cleaning and processing when porting across databases were also commonly cited, as well as a lack of effective, privacy-compliant data sharing methods to ensure data integrity. P03, a technician at a research center, explained her concerns with data security and transfer in her own work:

> *“The thing for me is that I don’t want someone to be able to go into my experiments folder… [I]f I share … a protocol with somebody and I send them a link from my network drive, I don’t want them to then be able to go into that and change it. So I will typically just e-mail people attachments because I like to keep raw versions in my folder that nobody else has touched. But yeah, it does get a little bit challenging when you’re sending massive data files to people.”* – (P03, research technician)

With respect to clinical research, participants described how a lack of unity across clinical trial management systems can lead to confusions when managing data. As P07 explains,

> *“Based on the study that we’re using, the portals could be different… the biggest issue with it is now you can have up to 12 different portals and 15 different logins… the amount of passwords and logins… it’s a headache. So the pharmaceutical companies … like go to third parties most of the time, and then some of them have their own portals… So then you have to use their portals… to then get the information back to the sponsor. So it’s all these separate branches that all then come together and you put that into the main one, which is the [electronic data capture system]… The hardest part is that there are so many different systems, like there isn’t one centralized one that everyone uses…. It is really trial- and sponsor-dependent – also system-dependent too.”* – (P07, clinical research assistant)

With respect to clinical data, participants mentioned the need to develop and incorporate tools that can handle electronic data capture (EDC) input errors. As P08 describes,

> *“There is a large amount of data coming from EHRs that is not well-structured. And how do you process that? So things like … a genetic test, or something that might be a scanned image… or unstructured clinical notes going through. So that’s an area where [we’re] sort of trying to figure out how we can enrich the data with some of that using things like ML and NLP … as part of our pipeline…. We’re dreaming for the point where we can actually use some of the interoperability standards, like FHIR, to get data directly. But most of the industry is sort of going into that kicking and screaming.”* – (P08, biotechnology research scientist)

In the handoff that occurs following data passing through Data Integrity stage, the quality-controlled, processed data are transferred on to additional professional stakeholders such as data scientists or bioinformaticians for data analysis. Challenges found in this transfer included ensuring data privacy and security, meeting regulatory requirements, learning diverse data storage and sharing systems, and supporting coordination among multiple stakeholders. P7, a clinical research assistant, elaborated on the current state of collaboration networks for clinical research:

> *“I would say with the collaborating side is that a lot of times, sponsors are really unrealistic with expectations and just goals overall. I think that they send like 5 emails in a day reminding you to put information in… and they want it in 3-day window after a patient comes in… But they won’t get back to you in a timely manner most of the time if you have a question about something or if you have an issue with anything… it’s kind of just an unfair relation[ship].* – (P07, clinical research assistant)

## 3. Data Analysis

The third theme that emerged was “Data Analysis”, which involves statistical analysis, machine learning modeling, and biological interpretation of results. Incorporation of public knowledge is often included, as well as integration of multiple data modalities. Tooling requirements of participants at the Data Analysis stage varied across research objectives. Particularly for participants with less experience in the field of computational biology, they described significant learning curves with regards to conducting computational analysis workflows for the first time.

> *“I think the biggest pain point was early on in getting started. I’m a biologist and a physician by training. I did not have [a] computational [or] coding background before diving into this work… And I think I … really would have benefited from … a more didactic or more guided kind of approach … to it, both as learning the actual coding … and then just learning some of the QC things to keep in mind. I ended up with a very convoluted workflow that I had created and coded myself.”* – (P11, clinician working in computational research).

P01, a statistical consultant for clinical trial analysis, expanded on the issues surrounding learning curves given the diverse skills and platforms needed to conduct biomedical research:

> *“One of the biggest funnels … comes down to potentially skill set and limited resources within particular organizations and systems. So finding someone who is strong with a particular system,… who can write code. Who can organize the data in such a way that it expedites the ability to move that through the data chain to get to … analysis [so], those data can be insightfully reacted upon.”* – (P01, statistical consultant)

Participants also mentioned a lack of standardized processes for version control of code and data. Participants working in computational biology research described how they needed to use both Python and R environments for their analysis work, and that continually transitioning back and forth between the two platforms was often an ordeal. Interviewees working specifically with large-scale ‘omics data described how the scale of their data can make analysis and debugging in local environments infeasible.

> *“[Be]cause of the scale of the data… you’re stuck running things in the cloud… I think the biggest problems that I’ve seen have to do with just debugging. It’s just an endless, endlessly frustrating process … because … you have to … log into the [virtual machine] and then get into the container and then try and figure out … what your commands are doing. So that would be my biggest frustration in general with … any of these workflow management systems. You can kind of shrink it. You can download all the containers … onto your desktop, … but it takes forever and you … can’t really run anything beyond trivial datasets. …Things are always different when you scale up to actual data set sizes…. There’s always a scaling issue. You could have a couple of reads, which could … somehow wreak havoc on your entire process.”* – (P15, bioinformatician in industry)

Both the diversity of coding environments and software and the lack of effective methods for multiomics data integration hamper research participants ability to conduct reproducible analysis, adding to the time required at the Data Analysis stage. P10 elaborates on issues regarding version control of software in the genomics field:

> *“There [are] some algorithms that go all the way back to using 2.7 Python, and you’re using … [version] 3.8. And because there [are] so many tools being developed, it’s usually really hard to pinpoint, all right, what version do I need? How’s the best way to make this environment, which is probably why pipeline software has become more popular and trending is because that is a massive issue for everybody.”* – (P10, bioinformatician at a research university)

In the handoff that takes place after Data Analysis, robust findings are passed on to stakeholders for further interpretation. This handoff involves collaboration between research scientists, clinicians, and relevant stakeholders and challenges found include lack of standardization in version control expectations for code and data, and difficulties meeting regulatory expectations, bottlenecking because of a need for expert insight and interpretation, and latency facilitating communication among data generators and research scientists involved in data analysis and the hand-off interpretation.

## 4. Data-Driven Discovery

The fourth theme of biomedical discovery research identified from our interviews was the stage of “Data-Driven Discovery.” Here, communication with collaborators and publication of manuscripts and data or workflows are used to advance knowledge in the field and disseminate findings. At this stage participants conduct a cycle of validation and verification to ensure research findings match regulatory expectations. Challenges found among participants during the Data-Driven Discovery stage included proper biological interpretation of results, meeting required regulatory expectations, and appropriately conveying the significance of conclusions drawn to public audiences.

Ultimately, across the themes identified for biomedical discovery, participant interviews all echoed a single message: the significance of collaboration and trust surrounding the data. As P10 explains,

> *“There [are] some groups that are definitely interested in biology and then there [are] other groups that are more [focused on] informatics work. Usually if there’s a good collaboration, [the biologists] trust [the bioinformaticians] to take care of all the smaller things like script management and trust in [our] overall analysis.* – (P10, bioinformatician at a research university)

Each exchange of data involved multiple professional stakeholders, including data generators, research scientists, data curators, third-party vendors, bioinformaticians, computational biologists, biologists, and clinicians. In many cases, data handoffs were not straightforward and involved a back-and-forth communication circle. For work intended to proceed toward human studies, FDA compliance was found to be required, and in all cases, insight and interpretation was continually needed from all stakeholders involved to ensure the accuracy and integrity of the data.

### A Proposed Framework for Biomedical Discovery

Informed by the general needs and challenges derived from half of the interviews and the themes derived from the second half of participant interviews, a biomedical discovery process emerged from the qualitative data analysis agnostic of data modality under consideration across participants (Figure 3).

**Figure 3.**
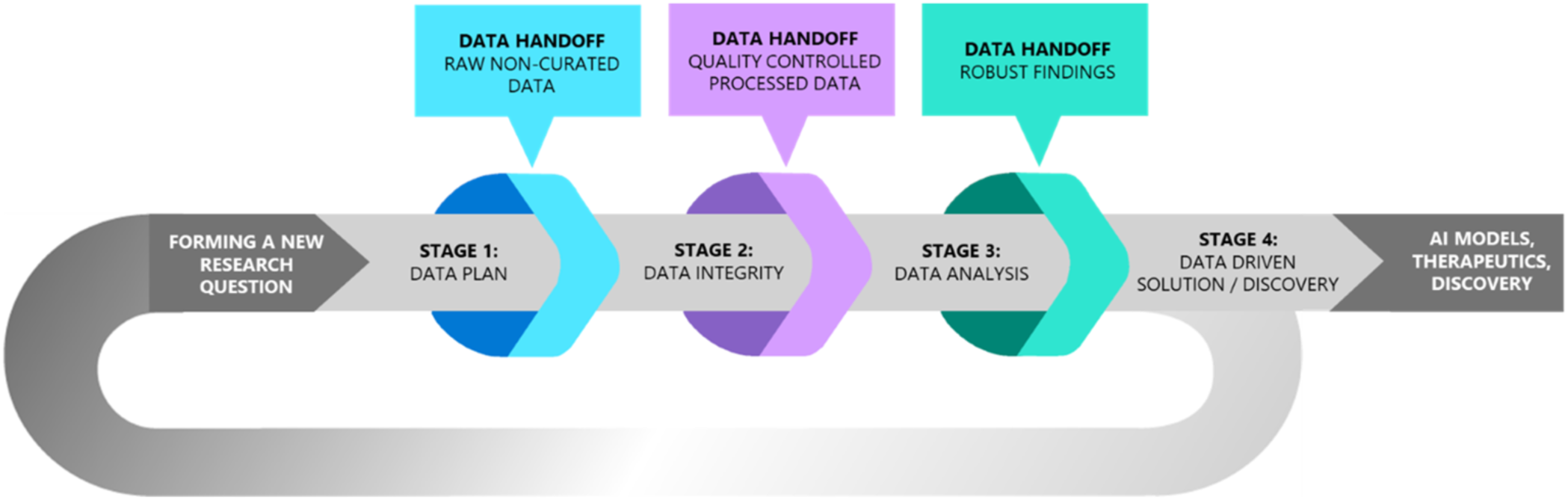
Visualization of the process of biomedical discovery.

Based on the data analysis four consecutive “stages” of biomedical discovery themes were identified, (1) Data Plan, (2) Data Integrity, (3) Data Analysis, and (4) Data-Driven Discovery, each serve as a distinct stage. Between each stage, a hand-off of data takes place among professionals involved in the biomedical research process. The formation of a research question to address a gap in the literature or fulfill an unmet clinical need is the start of the process, which leads into the first stage of research, the Data Plan. The results of the final stage, Data-Driven Discovery, funnel either into a new solution in the field or return to the start of the process to raise a new research question. Data can flow linearly through the process or return to previous stages depending on the needs of the research team. Table 1 provides a detailed overview of the required tasks and data jobs-to-be-done across each stage.

**Table 1.**
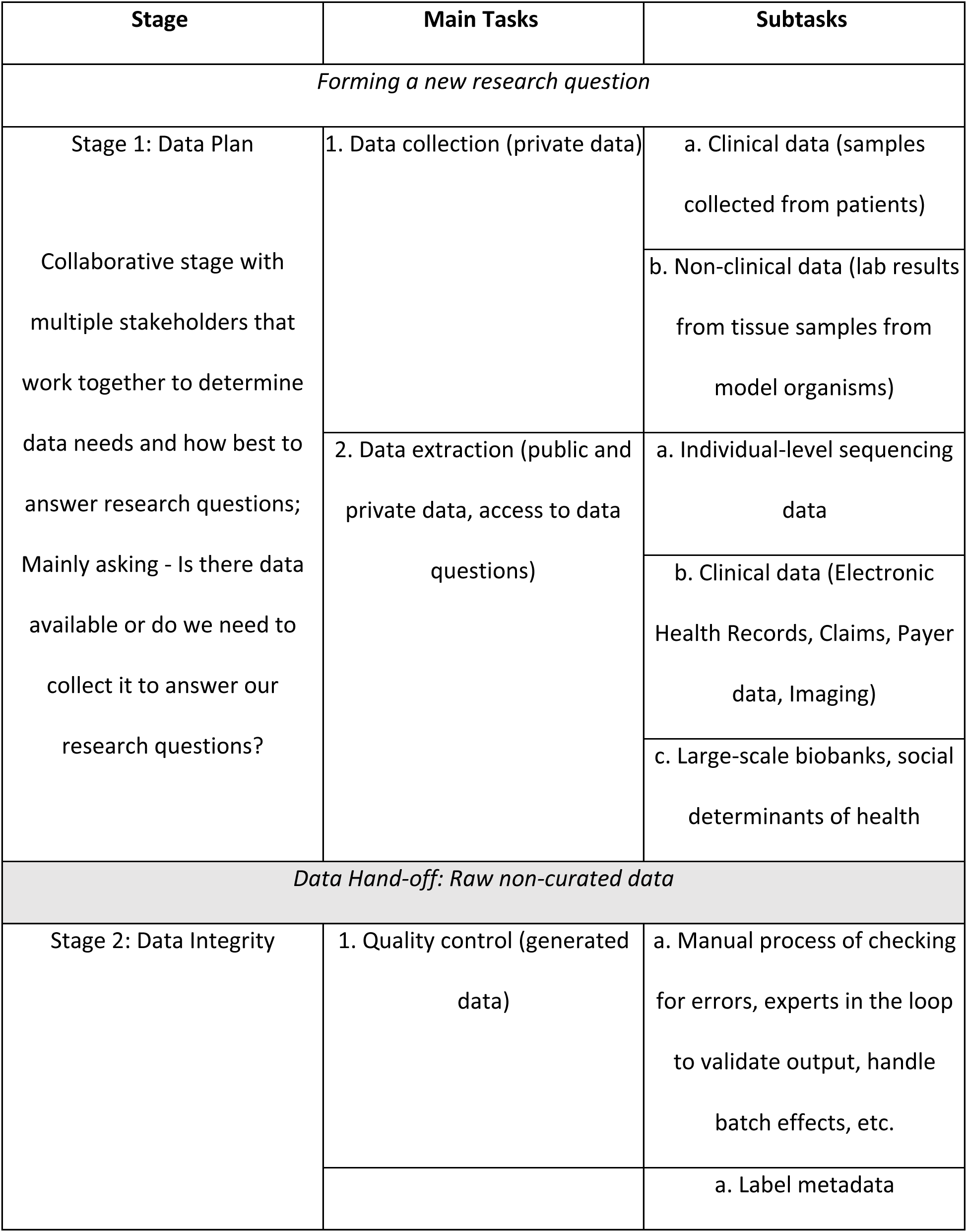
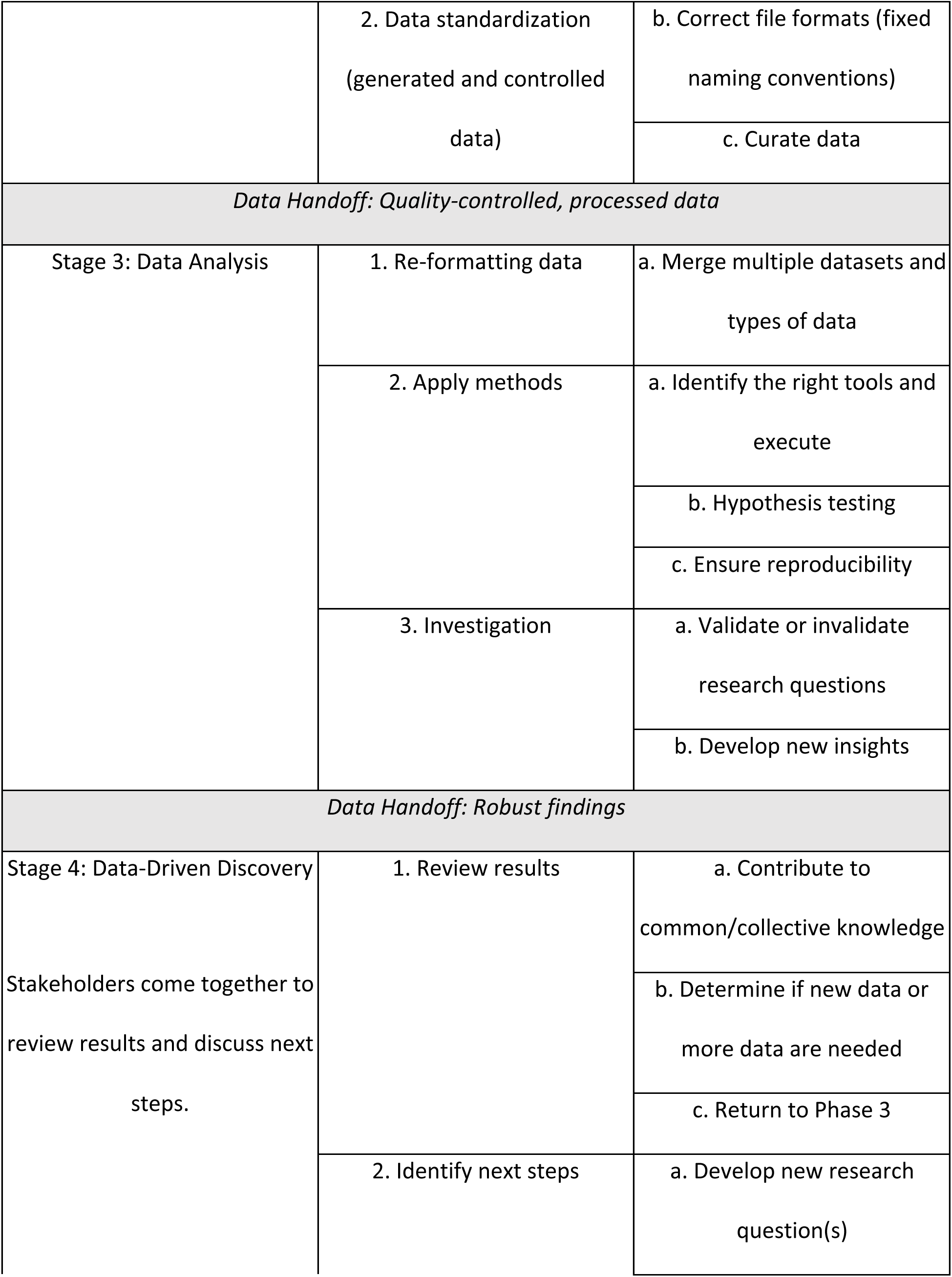
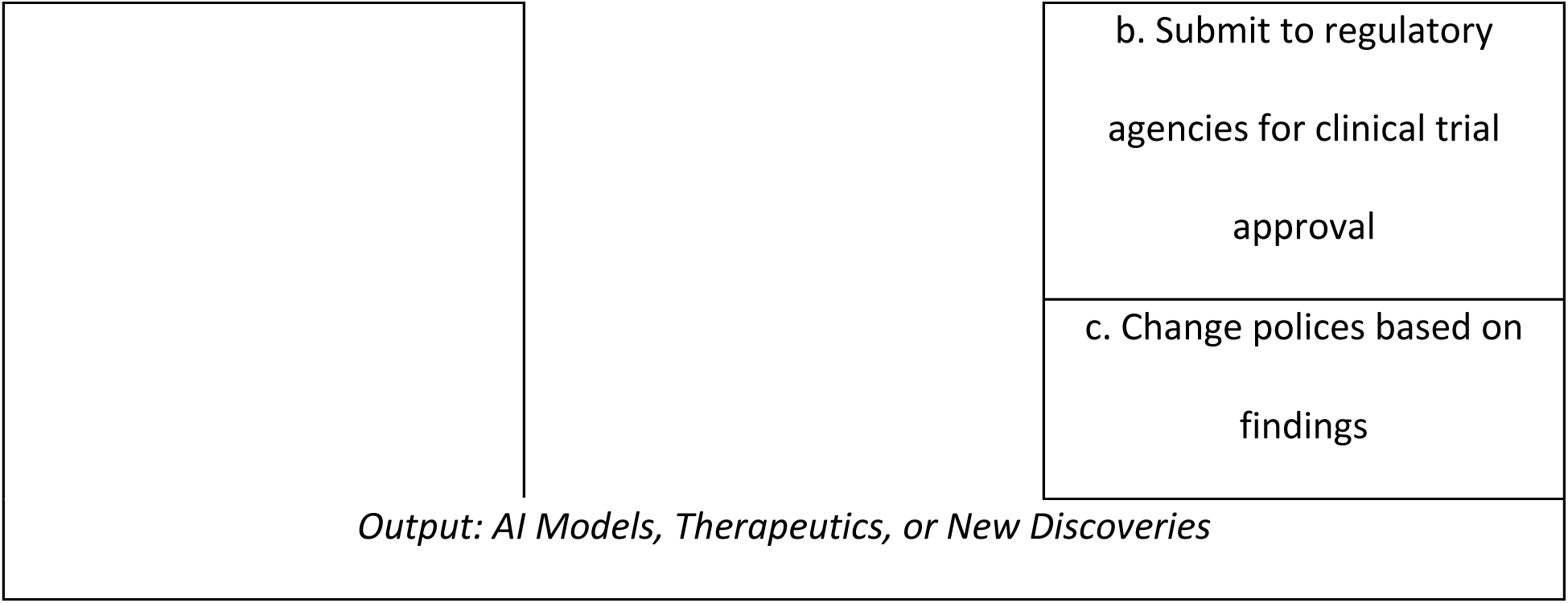
Overview of tasks and data jobs-to-be-done by stage.

### Levels of Biomedical Discovery

Given the various professional stakeholder roles and data modalities analyzed in our study, three subthemes emerged from the study data, each data task described by participants fit into three “levels” of discovery – (A) Non-human discovery, (B) Non-clinical human discovery, and (C) Clinical human discovery. The entire scope of biomedical research can be thought of as a funnel, with the range of translational implications starting broad in Level A (Figure 4). As time progresses, research evidence narrows to an individual translational model or therapeutic product.

**Figure 4.**
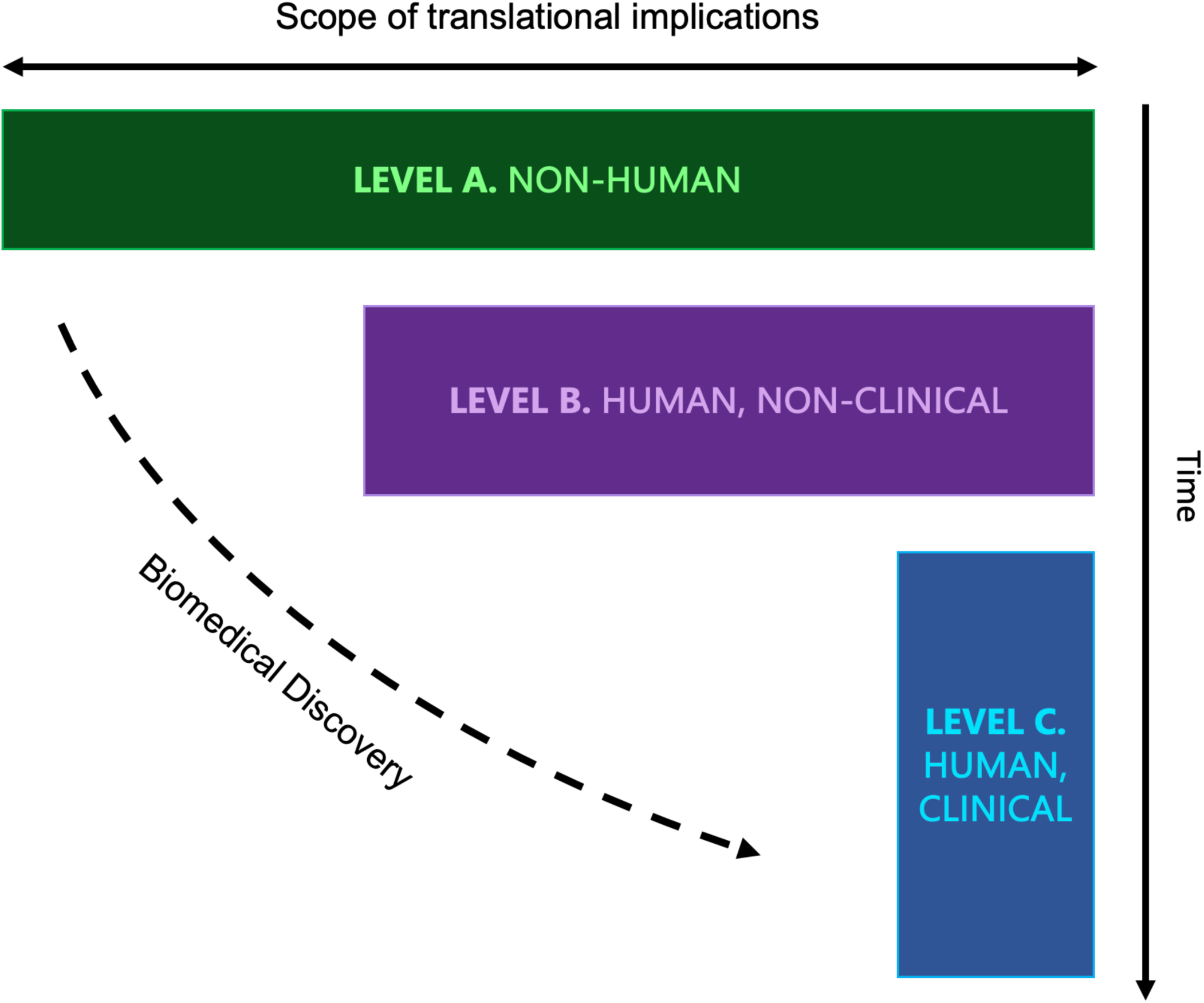
Visualization of the levels of biomedical discovery.

Within each level, researchers iterate through the biomedical discovery process as research questions are addressed and new questions emerge. The key insight from this movement of discovery is that the translational findings that directly impact precision medicine have their core roots in basic biological research. As P08 summarizes,

> *“How do we get this incredibly complex multidimensional data in a format that’s usable for the [computational biology] people to … [gain] insights[?] [S]o if you’re thinking about … software development, we [a]re really trying to take … a vertical slice through the whole stack and get everyone working together rather than having … the PhD scientists throwing it over the wall to the computational biologist who then [need] us to throw the data over the wall to them, like, really try to close that gap and get the teams working on a task much more closely aligned… You’ve got completely different needs if it’s … in vitro or in vivo. But the way you’re collecting data and analyzing it isn’t necessarily different.”* – (P08, biotechnology research scientist)

Figure 5 presents the full proposed framework of biomedical discovery research, including an itemization of tasks according to both level and stage of research.

**Figure 5.**
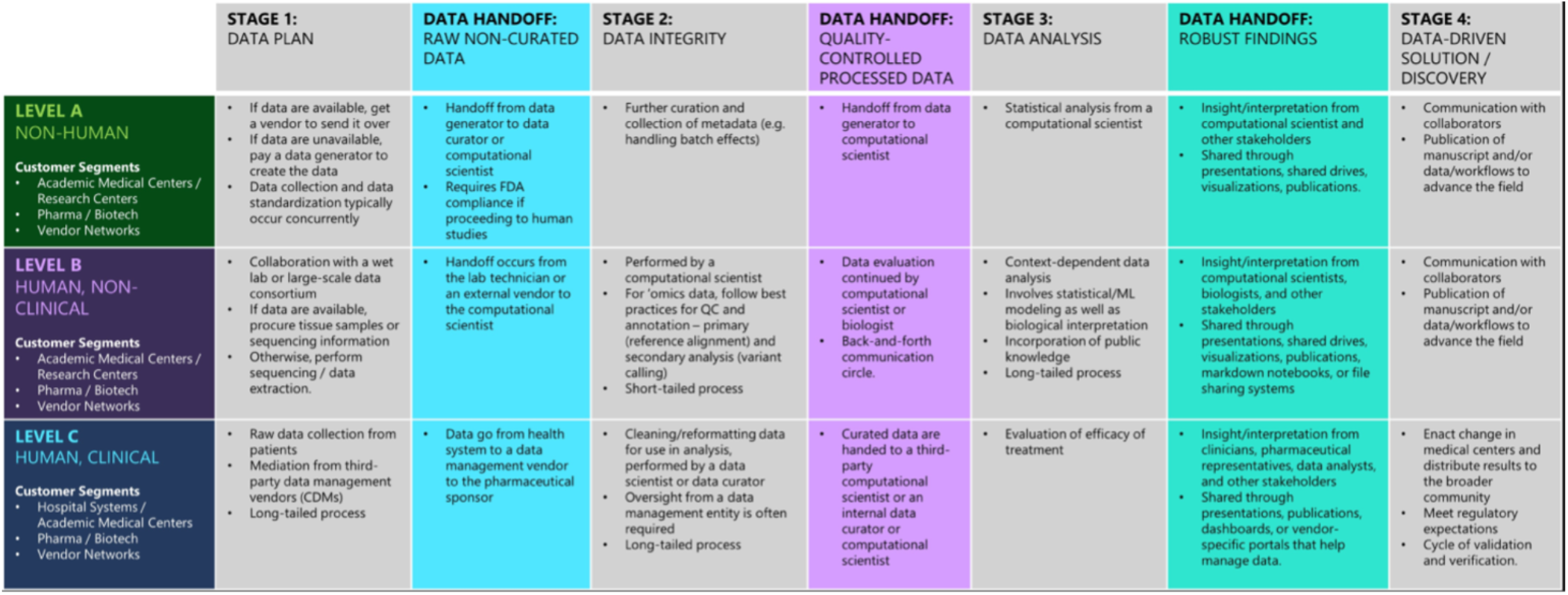
Biomedical Discovery Framework: A breakdown of tasks within each stage and each level of biomedical discovery.

## DISCUSSION

Data integrity and interoperability are essential to improving our ability to achieve precision medicine. However, most of the research conducted today is fixated on the development of new tools and methods for analysis. This myopic focus ignores the gaps in biomedical experimentation that lead to failings of data interoperability. To identify these omissions, our work aimed to identify and define a standard process of biomedical discovery research across professional stakeholder roles, research goals, and data subtypes. Our qualitative study provides insights into the data journey across stakeholders involved in biomedical discovery. We conclude our Discussion with a summary of key opportunities and future directions for those interested in addressing open challenges in the discipline.

Our study findings confirm many of the data challenges found in the literature with respect to biomedical discovery research, such as concerns related to (1) secure data storage, warehousing, withdrawal, access, and sharing, (2) quality control and curation of unstructured data, (3) processing and multiomics integration for large-scale, heterogeneous data, (4) reproducibility and version control, and (5) coordination and collaboration among stakeholders. In addition to challenges around regulatory standards and collaborative data partnerships. Finally, we found the importance of data integrity in hand-offs between the stages of biomedical research, and how data integrity is often assumed by data professionals who do not collect and curate their own data, due to the isolation of their work from other steps in the research process ecosystem. The proposed framework for biomedical discovery could help establish a common process among researchers to help address some of the biomedical data challenges as whole community to advance precision medicine research.

There are other proposed frameworks within the realm of biomedical discovery (Supplemental Table 1) but each only addresses a specific research context related to tooling needs and data analysis and presumes quality and integrity of the data. None of these frameworks capture the full scale of biomedical discovery across data modalities and stakeholder roles, while also considering the scope of data interoperability and integrity. One notable example of a framework that has successfully reduced the time spent on research development is the drug discovery process, however, this process is specific to drug development and does not include other therapeutic or AI precision medicine discoveries. Our proposed process allows us to reframe the entirety of biomedical discovery from the purview of foundational research, and by demystifying this step of work, we offer a clear opportunity to quantify the breadth of research that contributes to precision medicine.

We break the process of biomedical investigation into three key levels: non-human work, non-clinical human work, and clinical human work. We found a similar cyclical template of data planning, data integrity, data analysis, and data-driven discovery across all levels of biomedical discovery. Through the proposed framework, we identified the ability to categorize both the type of research being conducted as well as the step in the process to which it corresponds may help reduce the turn-around time of knowledge discovery. The proposed process emphasizes the need for secure collaboration and data analysis, with a focus on reducing data handoff miscommunication, early-stage data extraction errors, metadata errors, and reformatting errors during analysis to meet compliance and regulatory standards. Additionally, it can provide guidelines for how participants should standardize each step of analysis to expedite different stages of research. Regulatory bodies such as the FDA could use such a flow to clarify their expectations regarding biomedical research, facilitating a simplified submission process for research groups and a more thorough cycle of data validation and verification. The proposed framework could help foster data interoperability across the landscape of biomedical investigation through its definition of a unified procedure for research.

Many questions have been raised in regard to the implications of generative learning technologies such as GPT for biomedical discovery.[38] The proposed framework in this manuscript can be used in complement with the abilities of generative learning methods to facilitate the development of new knowledge – by incorporating the guidelines established in our process, generative learning environments could become a tool to enable streamlined research. For instance, users might describe their research focus and the stage of their work and ask for suggestions regarding how to accelerate their work. The generative learning software would then use the guidelines established in our biomedical discovery process to suggest possible research directions, analyses, and project management strategies. In the future, such framework-guided models could assist with building patient cohorts for research and clinical trial matching[39], standardizing and curating raw data, collecting metadata for data handoffs and changes to facilitate regulatory compliance, automating integration of heterogeneous data, and identifying the most suitable data for the chosen research question.

Our study was limited by the logistics of recruiting participants, as it had to be conducted virtually. As a result, future work is needed to validate the proposed framework with additional participants. Furthermore, the proposed process must be applied in real-world research projects and evaluated for its ability to improve biomedical discovery research. Another future direction of this work involves additional investigation of the third-party data management and contract research organization vendor networks to gain a full grasp of the data exchanges that take place in large-scale biomedical discovery projects.

### Key Opportunities for Biomedical Discovery

Based upon our data analysis findings, several key opportunities were identified for organizations looking to enhance their ability to conduct biomedical discovery research:

*(1) A user-friendly platform for bench-side data collection in biological research*. A transition from manual to electronic data collection in biologic discovery would increase efficiency, improve trust in the data collection and data analysis process for bench-side scientists, and improve interplay between wet and dry lab research.
*(2) A unified system for reproducible research.* An example of a group implementing such a system is the single-cell community, which consistently makes use of the Seurat and Monocle packages for its research. A unified system for data analysis would allow for consistent, sharable workflows and lead to a lower barrier to entry for computational analysis.
*(3) A simplified workflow for debugging and integration from notebooks into workflows to handle the large scale of ‘omics data.* This workflow may include the option to version control markdown documents and notebooks, as well as a graphical user interface to facilitate debugging in the cloud.
*(4) The study of third-party data management vendor networks for drug development.* Currently, the robustness of the IT infrastructure for a project can vary extensively depending on the organization in charge – larger companies tend to have stronger, cloud-based infrastructures for data storage and administration. More data mean more complications in terms of data processing, data transfer, and analysis, and in such situations, multiple experts from a variety of fields are required to manage the data. Third-party data management vendors are highly useful in managing these data access issues as well as facilitating regulatory proceedings for pharmaceutical companies. A better understanding of the systematized data exchange that occurs across these would vastly expedite biomedical discovery.
*(5) Improved tooling for clinical trial ingestion*. Multiple opportunities lie in the ability to handle free-form text to minimize data loss, the integration of NLP and ML, and education for clinical data collectors in the recording of patient data.
*(6) Improved communication between clinical trial managers and clinicians*. The wide variation in the data sharing systems that are used across pharmaceutical companies and third-party vendors results in a tremendous burden on clinical trial facilitators to ensure the ongoing viability of the trial. Multiple data management portals may be required for a project depending on the type of data being used. Clinicians are also often not able to directly see the impact of the work they help facilitate. We can reduce the turn-around time for biomedical discovery in the clinical space only through effective collaboration and communication between clinical trial managers and clinicians.
*(7) Integration of the framework into artificial generative intelligence (AGI) to accelerate research*. By incorporating the guidelines established in our biomedical discovery process into generative learning models such as GPT[38, 39], future researchers can expedite the pace of their research by receiving suggestions for how to unify their workstreams to others in the field, promote interoperability in their work, and identify the best data and the best tooling for their research goals.

## CONCLUSION

In this study, we identified and defined a process for biomedical discovery research lifecycle. We proposed a framework for research that tracks the movement of data across stages and levels of biomedical discovery that aims to unify biomedical stakeholders and align workstreams across research groups and organizations for improved data collaboration and integrity. We demonstrate how a proposed framework could help address some of the data and collaboration challenges to improve data integrity and interoperability for knowledge discovery. A standard process is particularly crucial to ensure the accuracy and reproducibility of biomedical models when considering transitions toward production-level applications in healthcare and the life sciences.

We also made several suggestions for approaches to accelerate biomedical discovery, including cloud-based computational infrastructures for centralized data warehousing and withdrawal, improved debugging workflows for the analysis of large-scale, heterogeneous data, new methods for the ingestion of unstructured data, and the establishment of vendor networks to facilitate data management and the fulfilment of regulatory requirements. In particular, we see the relevance of incorporating a proposed framework into future artificial generative intelligence platforms.[38] In concert with access to data resources and regulated data curation and analysis methods, an augmented AGI platform could provide researchers with assistance in hypothesis generation, data harmonization, exploratory data analysis and visualization, data processing and curation, and the proposal of relevant datasets and methods. Such a system could identify the most appropriate data for a given research aim, ensure the fulfillment of guidelines during quality control and analysis, and assist with reporting the results of a study and prompting future research directions.[39]

More research is needed to validate if such a proposed framework could improve large-scale multiomics data integrity, interoperability, analysis, and collaboration challenges. Through its application, we hope to see a paradigm shift in biomedical discovery research practices, bringing us exponentially closer to realizing individualized therapeutics for all patients and fulfilling the promise of precision medicine.

## Data Availability

All data produced in the present study are available upon reasonable request to the authors.

## ACKNOWLEDGEMENTS

We would like to thank Odeline Mateu-Silvernail for her help with the development of the visual representation for the final biomedical discovery process. We would also like to thank Erdal Cosgun, Mamta Giri, Roberto Lleras, Venkat Malladi, Chuan Li, Chaitanya Bangur, Jeremiah Wander, and Scott Saponas for their feedback and support throughout this study.

