## Supplemental Tables for "Accelerating precision medicine: a proposed framework for large-scale multiomics data integrity, interoperability, analysis, and collaboration in biomedical discovery"

### SUPPLEMENTAL DATA

Supplementary Table 1. A list of all identified frameworks in the healthcare space

| Paper | Publication / Conference | Year | DOI | Data types considered | Personas considered | Scope of framework | Summary |
|---|---|---|---|---|---|---|---|
| **A framework for big data technology in health and healthcare[40]** | 2017 IEEE 8th Annual Ubiquitous Computing, Electronics and Mobile Communication Conference | 2017 | 10.1109/UEMCON.2017.8249095 | Healthcare data (healthcare provider data, EMRs, insurance company/payer data, patient data, wearables) | N/A | Clinical research | Summarize options for clinical data sources, big data storage and analysis systems, and translational opportunities for clinical data in a 4-step process |
| **A framework for the use of genomics data at the EPA[41]** | Nature Biotechnology | 2006 | 10.1038/nbt0906-1108 | Genomic data | N/A | Human non-clinical research and disease diagnosis | Set of guidelines to be considered when working with genetic data |
| **A Harmonized Data Quality Assessment Terminology and Framework for the Secondary Use of Electronic Health Record Data[42]** | eGEMS (Generating Evidence & Methods to improve patient outcomes) | 2016 | 10.13063/2327-9214.1244 | Clinical data from EHRs | N/A | Clinical research | Multiple studies evaluating data quality in clinical research were harmonized to construct a unified set of requirements |
| **An Integrated Data Management Framework for Drug Discovery – From Data Capturing to Decision Support[43]** | Current Topics in Medicinal Chemistry | 2012 | 10.2174/156802612800672862 | Chemical data related to drug discovery and development | N/A | Drug Discovery | Drug discovery informatics platform that allows for management of multiple reagents compounds, and assays |
| **Argonaut: A Web Platform for Collaborative Multiomic Data Visualization and Exploration[44]** | Patterns | 2020 | 10.1016/j.patter.2020.100122 | Multiomics data | N/A | Data visualization and analysis for multiomics research | Secure, web-based sharing of data analysis and visualization for multiomics data |
| **Assuring the Machine Learning Lifecycle: Desiderata, Methods, and Challenges[45]** | ACM Computing Surveys | 2021 | 10.1145/3453444 | Data agnostic | N/A | Machine Learning Analysis | Defined a 4-step process / iterative loop for the lifecycle of machine learning analysis |
| **Best practice data life cycle approaches for the life sciences[46]** | F1000Research | 2018 | 10.12688/f1000research.12344.2 | Genetic sequencing and annotations, metabolite/proteomic profiles | Research scientists | Life sciences / Biomedical / Biosciences / Bioinformatics Research | Best-practice lifecycle for life sciences / biomedical data, highlighting data reusability |
| **Best practice framework for Patient and Public Involvement (PPI) in collaborative data analysis of qualitative mental health research: methodology development and refinement[47]** | BMC Psychiatry | 2018 | 10.1186/s12888-018-1794-8 | Qualitative mental health data | Patient and Public Involvement (PPI) Researchers | Mental Health research | Multi-stage framework for qualitative research in the mental health space |
| **Building Highly-Optimized, Low-Latency Pipelines for Genomic Data Analysis[48]** | Conference on Innovative Data Systems Research | 2015 | N/A | Genomic Sequencing data | N/A | Aligned/annotated genetic read analysis for association and causality | Multi-step pipeline for QC and analysis of genomic sequencing data |

| | | | | | | | |
|---|---|---|---|---|---|---|---|
| **Clinical data quality: a data life cycle perspective[49]** | Biostatistics & Epidemiology | 2020 | 10.1080/24709360.2019.1572344 | Clinical data | N/A | Clinical research, clinical trial. Recruitment, phenotype-driven rare/genetic disease research, large-scale observational studies | Multi-stage data life cycle for clinical data quality, use, and reuse |
| **Cloud-based Healthcare data management Framework[50]** | KSII Transactions on Internet and Information systems | 2020 | 10.3837/tiis.2020.03.006 | Clinical data (from unstructured patient data to EHRs) from healthcare organizations | N/A | Cloud-based management and analysis of healthcare data | Cloud Infrastructure for the ingestion, storage, and processing of clinical data |
| **Defining and Developing a Generic Framework for Monitoring Data Quality in Clinical Research[51]** | AMIA Annual Symposium Proceedings | 2018 | N/A | Clinical data | N/A | Evaluate quality of data in clinical research using this framework | A "Fit-for-use" data quality monitoring framework, presented as a nested concentric network, can facilitate increased efficiency in clinical data quality monitoring |
| **Development of a Genomic Data Flow Framework: Results of a Survey Administered to NIH-NHGRI IGNITE and eMERGE Consortia Participants[52]** | AMIA Annual Symposium Proceedings | 2019 | N/A | Genomic data | Patients, Genomic researchers, Clinicians | Interpretation of genetic profiles in clinical settings | A mapped exploration of data processing and analysis for clinical interpretation of patient genetic information |
| **Ethics of Using and Sharing Clinical Imaging Data for Artificial Intelligence: A Proposed Framework[53]** | Radiology | 2020 | 10.1148/radiol.2020192536 | Patient data | Patients, Researchers, Clinicians, Administrators, Payers, Purchasers, Industry | Secondary use of data (research) requires researchers to act as ethical data stewards | Set of ethical considerations as. Medical imaging data are used for research |
| **Gathering and Learning from Relevant Clinical Data: A New Framework[54]** | Academic Medicine | 2015 | 10.1097/ACM.0000000000000508 | Clinical data | N/A | Standardize clinical practice | Theory-built framework called the Standardized Clinical Assessment and Management Plan (SCAMP) |
| **Impact of a five-dimensional framework on R&D productivity at AstraZeneca[55]** | Nature Reviews Drug Discovery | 2018 | 10.1038/nrd.2017.244 | Metabolite/pharmacological data | N/A | Pharmaceutical research / drug discovery | 5R framework (Right target, tissue, safety, patient, and commercial potential) can accelerate the process of drug discovery for big pharma |
| **Preparing Medical Imaging Data for Machine Learning[56]** | Radiology | 2020 | 10.1148/radiol.2020192224 | Radiology / Patient Imaging Data | AI researchers | Making medical image data available for ML/AI research into improving radiology diagnosis | Defined steps for process of medical image data handling |
| **Security model for Big Healthcare Data Lifecycle[57]** | Procedia Computer Science | 2018 | 10.1016/j.procs.2018.10.199 | Clinical data (from unstructured patient data to EHRs) from healthcare organizations | N/A | Identify weak security points in the lifecycle of healthcare data | Multi-stage data lifecycle going from data collection to knowledge creation for healthcare research using big data |

| | | | | | | | |
|---|---|---|---|---|---|---|---|
| **State of the Field in Multi-Omics Research: From Computational Needs to Data Mining and Sharing[29]** | Frontiers in Genetics | 2020 | 10.3389/fgene.2020.610798 | Omics data – genomics, epigenomics, transcriptomics, proteomics, metagenomics, etc. | N/A | Non-clinical research – defining best practices for FAIR sharing | A flow diagram defining best steps for data analysis, sharing, and reproducibility in a FAIR ecosystem |
| **Translational Research 2.0: a framework for accelerating collaborative discovery[58]** | Personalized Medicine | 2014 | 10.2217/pme.14.15 | Life sciences / biomedical data | Medical / Life science researchers, care delivery organizations | Represent the overarching ecosystem of biomedical and life sciences research | A depiction of the translational sciences ecosystem |

Supplementary Table 2. Data Types used by Participants

| **Data Types Used** |
|---|
| • ELISA / FISH / Flow cytometry data from model organisms and patient tissue samples |
| • Clinical data from patient electronic health records<br>○ Lab measurements, vital readings, biomarker/metabolite measures, imaging/radiology data, qualitative measurements |
| • Genomic data<br>○ Single-cell RNA-sequencing (scRNA-seq)<br>○ Whole Genome DNA-sequencing (WGS) |
| • Post-clinical data<br>○ Drug performance data<br>○ Drug marketing data |

Supplementary Table 3. Participant Methods for storing data

| **Methods for Storing Data** |
|---|
| • Microsoft Excel, Box cloud storage, SharePoint, OneDrive |
| • Watson LIMS |
| • On-premises clusters |
| • Epic/Epic CareConnect |
| • REDCap |
| • Benchling |
| • Amazon Web Services (AWS) RedShift/S3 |
| • Microsoft SQL Server |
| • Snowflake |
| • Google Cloud Platform (GCP) |
| • Third-party data vendors / EDC vendor-based portals<br>○ Medidata RAVE, Inform, IBM Clinical, Open Clinica, Triad<br>○ FlatIron, Syapse, Coda, IDT, GenScript |

Supplementary Table 4. Participant Methods for sharing data

| **Methods for Sharing Data** |
|---|
| • Presentations, slide decks with visualizations and data tables |
| • Data Visualization Platforms such as PowerBI and Tableau |
| • Markdown notebooks |

| |
|---|
| ○ RMarkdown for R<br>○ Jupyter for Python |
| • Shared drives, email, Slack, USB drives |
| • HIPAA-compliant Box storage |
| • Mirth HL7 Engine |
| • Cloud-based platforms (e.g. AWS Deequ) |

Supplementary Table 5. Participant Methods for handling data access

| **Methods for Handling Data Access** |
|---|
| • Internal IT departments<br>○ Linux chmod |
| • Role-based permissioning<br>○ Snowflake, AWS |
| • Third-party data management vendors |
| • Institution- or consortium-wide data governance councils |

Supplementary Table 6. Tools Used by Participants

| **Tools Used** |
|---|
| • User-friendly data analysis<br>○ IBM SPSS, REDCap, Epic, Excel/Macros |
| • Image analysis<br>○ ImageJ, Prism |
| • Evaluating data quality<br>○ AWS Deequ |
| • scRNA-seq<br>○ Seurat, Monocle, ScanPy |
| • Primary and secondary genomic data analysis<br>○ GATK: best practice workflow |
| • Tertiary / General Data Analysis<br>○ Python (pandas, numpy, scipy, Jupyter notebooks, VSCode)<br>○ R (Bioconductor, ggplot, tidyverse, RMarkdown, RStudio)<br>○ SQL<br>○ SAS |
| • Genomics-specific analysis<br>○ Google Genomics Pipelines API on GCP<br>○ Glow genomics toolkit on AWS Spark |
| • Pipelining and workflow development<br>○ Nextflow, Cromwell |
| • Heavy data analysis<br>○ RAPIDS framework, Vaex.io, IPyWidgets, CUDA Python (CuPy) |
| • Versioning<br>○ Anaconda, Docker |
